# Why simple face masks are unexpectedly efficient in reducing viral aerosol transmissions

**DOI:** 10.1101/2020.12.03.20243063

**Authors:** Steffen Freitag, Steven G. Howell, Kevin T. C. Jim

## Abstract

During the current pandemic and in the past, shortages of high quality respirators have forced people to protect themselves with homemade face masks that filter poorly in comparison to N95 respirators ^1–4^ and are often designed in ways that makes them susceptible to leaks ^5,6^. Nevertheless, there is compelling epidemiological ^7,8^ and laboratory evidence ^9–12^ that face masks can be effective in impeding the spread of respiratory viruses such as influenza and SARS-CoV-2. Here we show that this apparent inconsistency can be resolved with a simple face mask model that combines our filtration efficiency measurements of various mask materials with existing data on exhaled aerosol characteristics. By reanalyzing these data we are able to reconcile the vastly different aerosol size distributions reported ^13–19^ and derive representative volume distributions for speech and breath aerosol. Multiplying filtration efficiency by those aerosol volumes, which are proportional to emitted viral load, shows that electrostatically charged materials perform the best but that even most uncharged fabrics remove *>* 85 % of breath and *>* 99 % of speech aerosol volume for exhaled particles *<* 10 *µ*m in diameter. A leak model we develop shows the best uncharged fabric masks are made of highly air-permeable and often thin materials reducing viral load by up to 45 % and 50 % for breath and speech, respectively. Less permeable materials provide reduced protection because unfiltered air is forced through the leak. This can even render some charged materials inferior to uncharged household materials. Our model also shows that thin fabric masks provide protection for the wearer from aerosols expelled by another person reducing inhaled viral load by up to 20 % and 50 % and if leaks are avoided up to 35 % and 90 % for breath and speech, respectively.

---

Respiratory viruses such as influenza and SARS-CoV-2 (severe acute respiratory syndrome coronavirus 2) are detectable in the fluids of the upper respiratory tract of infected people ^9,20,21^ and can be expelled from the nasal and oral cavity within giant liquid aerosols, often referred to as droplets, that are formed when mucus and saliva layers break up during sneezing and coughing ^22,23^ and even speaking ^17,19^. The largest of these, with diameters *>* 100 *µ*m, rapidly settle to surfaces. Particles with diameters of a few tens of microns quickly lose water and shrink in relatively dry ambient air, so fall slowly and can remain airborne for extended periods. Even smaller aerosols are generated during these activities ^17,24,25^ that can stay airborne for hours but originate from deeper inside the respiratory tract.

Interest in these tiny aerosols, that are in fact the result of normal breathing, reaches back almost a century, but it was not until the late 1990s that exhaled breath aerosol was analyzed comprehensively with a laser-based optical particle counter ^13^. These observations have since been repeated with other measurement techniques yielding divergent results ^17^,19,24 but also reproduced multiple times ^14–16,18^, confirming the notion of two breath modes with average number concentration mode diameters around 0.95 *µ*m and 3.36 *µ*m at the high humidity inside the respiratory tract (see methods for derivation of mode diameters from existing data). The larger of these two modes originates from an area around the vocal cords (larynx mode) where air turbulence ruptures the laryngeal mucus layer during exhalation ^26^, while the smaller mode originates deep in the lungs. Studies show that these tiny aerosols form during inhalation when the lining fluid inside the lungs stretches and ruptures during reopening of the bronchioles that close during exhalation ^14,16,27^. A single study found an even smaller mode created in the lungs with a mean diameter near 0.15 *µ*m ^15^, which is supported by numerical modeling ^27^.

Face masks and respirators are used to prevent exposure to viruses transported on these aerosols, but may be beneficial if they merely reduce the viral load one is exposed to. It has been shown for influenza that, should infection occur, exposure to a lower viral load causes a less severe disease ^28^. To estimate viral load on expelled aerosols and potential reduction by face masks, we assume that viral concentration within the liquid particles is the same as in the respiratory fluid, so the number of viruses is proportional to particle volume. This assumption is necessary since there are no measurements of virus concentration per exhaled particle as a function of particle diameter. While different respiratory fluids likely also have varying virus concentrations, it appears that breath and speech aerosol volume are each dominated by single respiratory fluid sources (Extended Data Fig. 6). Thus, converting the existing number size distribution data to volume distributions (see methods) yields a size distribution of relative viral load. We further assume that these exhaled particle size distributions from healthy subjects are the same as those from infected individuals, as has been shown in a study on influenza virus ^29^.

## Filtration efficacy of various materials

Estimating the reduction of viral load for face masks requires information about the material’s filter penetration fraction (FPF), which describes filtration efficacy as function of aerosol size and flow rate. It is determined in the lab by challenging mask materials with laboratory-generated test aerosols at a range of flow speeds and subsequently dividing the aerosol concentration measured downstream by the concentration observed upstream of the material (see Eqs. 2 and 6 in methods).

Of the 20 materials tested in this study and summarized in Extended Data Table 1, two are cutouts from N95 respirators, 3M models 8210 and 8000. These respirator types filter aerosol at efficiencies of 95 % ^30^–33 or close to this value ^34^ at NIOSH (National Institute for Occupational Safety and Health) standard flow speed around their worst diameter, commonly referred to as most penetrating particle size (MPPS). For N95 respirators this size is 0.04 - 0.1 *µ*m in diameter ^30–34^, which agrees well with our results (Fig. 1a in blue and Extended Data Fig. 2 for the other N95 model). Higher velocity data show FPFs for both models considerably exceed 5 % (red in Fig. 1a) consistent with earlier observations ^35^.

**Fig. 1.**
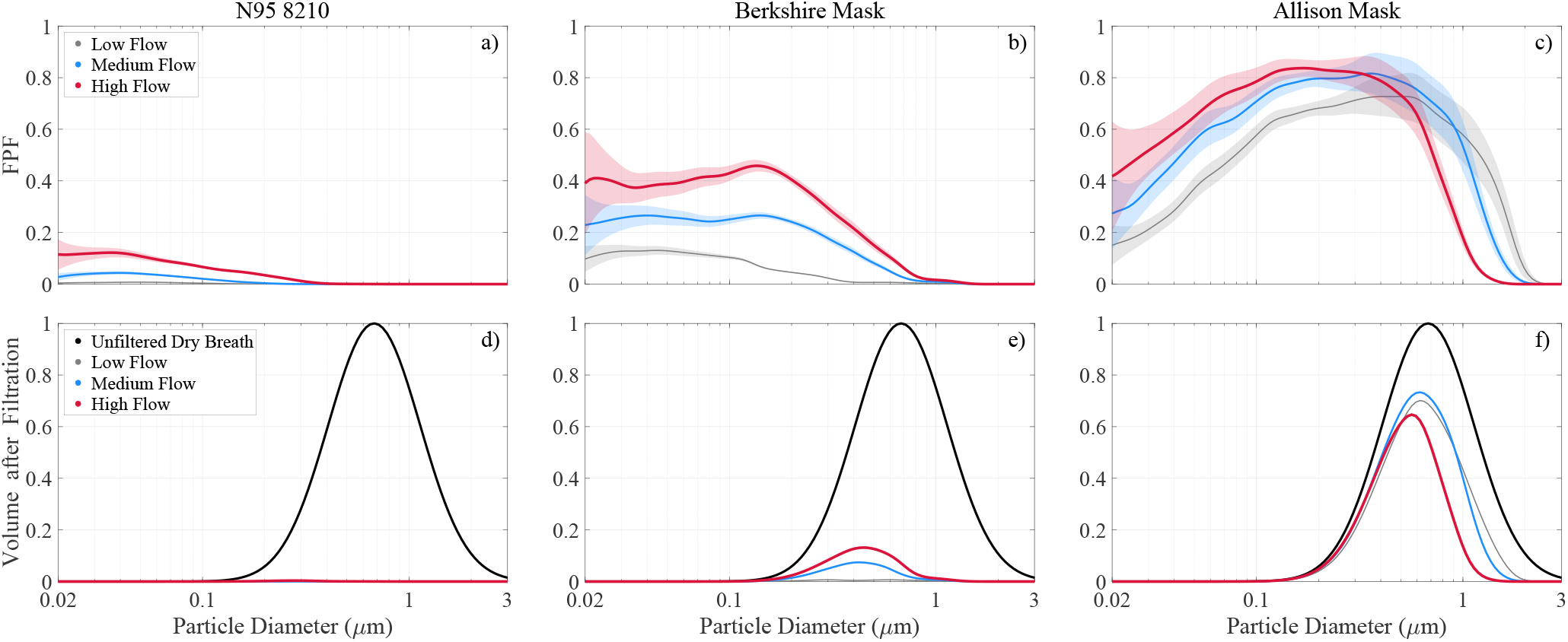
Filter penetration fraction and aerosol volume after filtration for selected face masks. (top) Measured FPF at low (2.7 cm s^*−*1^), medium (8 cm s^*−*1^) and1 high flow (22.8 cm s^*−*1^) in gray, blue and red, respectively. Shading illustrates measurement uncertainty (1*σ*) around the mean (solid lines). (bottom) Breath aerosol volume at 50 % RH before (in black; normalized to the maximum value) and after filtration at 3 selected flows (in gray, blue and red; same as top row). The Berkshire mask is a surgical-style mask designed for clean room use and includes an electret layer. The “Allison” mask has two knitted layers: the inner layer is 92 % rayon/ 8 % spandex, while the outer layer is a 400 g m^*−*2^ doubleknit Pont de Roma made of rayon, nylon, and spandex.

All filters capture aerosol mechanically by impaction and interception of larger particles that have too much inertia to avoid the fibers and by Brownian diffusion of very small particles, leaving particles in the middle, around 0.3 *µ*m, very difficult to capture. N95 respirators add an electret trapping mechanism that attracts aerosols of opposite charge (Coulomb force) and via charge separation in neutral particles (dielectric effect) using permanent electrostatic charges embedded in melt-blown polypropylene fibers during manufacturing. This dramatically improves FPF without increasing resistance to airflow ^36^. The Chinese KN95 masks also use electret material and show comparable FPF values (see Extended Data Fig. 2). However, one of the tested KN95 models is about two times harder to breathe through than N95 respirators (Extended Data Table 1). Note, the tested KN95-type models do not have the rigid hemispheric design of respirators so are referred to as masks here.

Homemade masks lack electret material, so have considerably worse FPF between 0.7 and 0.85 (Fig. 1c) and MPPS around 0.3 *µ*m. The other 8 tested household materials (shown in Extended Data Fig. 4g-o) exhibit comparable MPPS and FPF with the exception of the coffee filter and dust cover fabric, which perform worse. Our observations agree well with previous household material studies that reported FPFs around 60 - 90 % near their MPPS ^1–4^. As in earlier observations ^1^ we observe a sharp increase in filtration efficiency above 1 *µ*m with efficiency reaching 100 % near 3 *µ*m for all tested fabrics except the dust cover.

An electret surgical-style (Berkshire) mask we tested has FPF values between those for N95 respirators and household materials (Fig. 1b), because these masks are generally thinner and more permeable than N95 respirators (compare pressure drops in Extended Data Table 1) reducing time for particle trapping as air flows through the material. A lower fiber charge density also plays a role in its decreased filtration performance ^37,38^. Note that the wide variety of reported FPFs (5 - 95 % at MPPS of 0.04 - 0.35 *µ*m in diameter ^6,30,32,39^) indicates that some surgical-style masks lack an electret filtration layer.

Three electret air filters were examined as well, showing FPF distributions similar to the surgical-style mask because these materials are also designed for low air flow resistance ^33^ (3M Filtrete 1500 and 2500, Blue Air filter; see Extended Data Fig. 4). Of the materials only Filtrete 2500 outperforms the surgical-style mask at diameters above 0.1 *µ*m, the approximate size of influenza and SARS-CoV-2 viruses ^40,41^.

Halyard H600 is another electret ^42^ material tested (Extended Data Fig. 4c). It is used as surgical wrap in hospitals and was proposed by medical professionals as mask material earlier this year ^43^. The material consists of two easily separable layers (colored blue and white), which when tested have significantly different air flow resistance (Extended Data Table 1) but almost identical FPF, comparable to recent findings ^42^ and to the surgical-style mask although H600 is harder to breathe through.

## Viral load reduction of face masks

By reconciling existing data on exhaled aerosol characteristics we are able to derive representative speech and breath aerosol volume distributions. We model 4 scenarios that describe these distributions at two relative humidities (RH) (see Extended Data Fig. 6). The two “humid” scenarios represent exhalation through the face mask (protecting others) assuming a humidity of 99.5 % behind the mask, while the “dry” scenarios represent inhalation of particles through the mask that have shrunk in 50 % RH air after being exhaled by another person (self-protection). For breath aerosols, the volume-weighted geometric mean diameters for the scenarios are 0.68 (dry) and 2.15 *µ*m (humid), and the speech aerosol diameters are 2.75 and 8.59 *µ*m (Extended Data Table. 3).

Viral load reduction (VLR) is the fraction of virus (as represented by aerosol volume) removed by the mask. In the absence of leaks, viral load penetrating the mask for each of the 4 scenarios is just the integral of the volume size distribution and the FPF at a particular breathing rate, shown graphically for dry breath in the bottom row of Fig. 1. Unfortunately, as we see all around us, few masks actually fit tightly enough to seal, so some exploration of the effect of leaks is necessary. As a simple approximation for gaps on each side of the nose in the absence of a stiff nose bridge strip, we assume two leaks with a diameter of 1 cm each, comparable to that of a small finger of a human hand. Our model calculations also require a mask surface area *A*_M_ and breathing rate since FPF is a function of flow velocity. We use 189 cm^2^for the two N95 respirators ^44^ and 31 cm^2^ for all other face masks as well as a minute volume of 10 L min^*−*1^ (MV; inhaled air per minute) typical for breathing at rest ^45^ (see methods for details).

We limit our analysis to aerosol *<* 10 *µ*m in diameter because face masks with no leaks have a filtration efficiency of 100 % above 3 *µ*m as discussed above. When small leaks between face and mask are considered, some larger particles will slip through, but their number is likely to be small compared to those inertially impacted onto the mask because they can not make the sharp turns required to escape. We consider only breath and speech since coughing aerosols smaller than 10 *µ*m exhibit a volume distribution similar to that of speech ^17,25^.

VLR values for masks and the two respirators are shown in Fig. 2a-d where colors highlight material categories and darker and brighter shadings no-leak and leak conditions, respectively. The N95 respirators and KN95 masks (in red and yellow) have identical VLR signatures for all 4 scenarios because their VLR only depends on the amount of unfiltered air passing through the leaks (Fig. 2e) since the fraction of air that makes it through the mask has a VLR value of nearly 100 %. More importantly, these data show N95 respirators only reduce viral load by about 70 % with leaks present, merely 20 % more than the average VLR value for non-electret household materials (in brown and gray) except for the case of dry breathing. The household materials perform poorly in this case (VLR *<* 20 %) because most of the viral load is on particles below 3 *µ*m above which most materials show 100 % filtration efficiency (Fig. 1c and f).

**Fig. 2.**
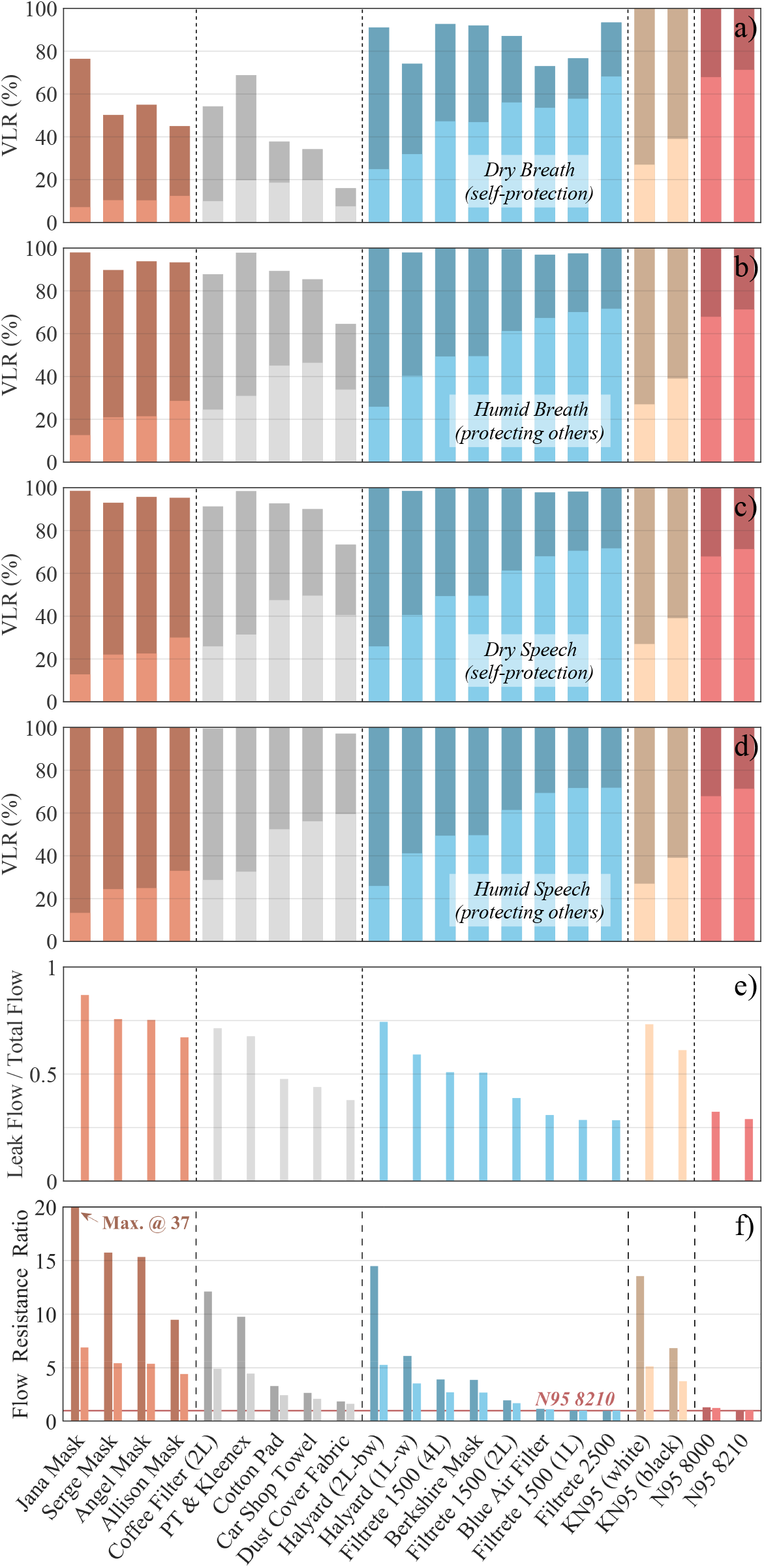
Overall protection value of tested face masks. Face masks are grouped by type: fabrics used in sewn masks (brown), non-woven household materials (gray), weakly electret filters (blue), KN95’s (yellow) and N95 respirators (red). Brighter and darker bar colors show performance with and without leaks. Top 4 graphs show viral load reduction (VLR) during (a) dry breath aerosol inhalation (at 50 % RH) and (b) humid exhalation (at 99.5 % RH) as well as (c) dry speech aerosol inhalation and (d) humid exhalation. Plot e) shows leak flow ratio and f) flow resistance in comparison to the N95 8210 respirator (red solid line in the background). Calculations are based on a minute volume of 10 L min^*−*1^ (breathing at rest) and a surface area of 189 cm^2^ for N95 respirators and 31 cm^2^ for all other face masks.

When leaks are included, the face masks provided by Hawaiian mask makers show the lowest VLR values of the tested materials (in brown in Fig. 2, 10 - 25 % VLR) with exception of the “Allison” mask, even though their FPF distributions are comparable to the other household materials, because these masks are made out of multiple fabric layers or very tightly woven cotton (Extended Data Table 1) reducing the mask’s permeability and consequently forcing unfiltered air through the leaks instead (Fig. 2e). Their low permeability also makes them about 5 times harder to breath through than a N95 respirator (Fig. 2f). When properly sealed, these masks provide exceptional protection with VLR values *>* 90 % in all cases except dry breath but are 15 - 37 times harder to breathe through than a N95 respirator decreasing chances of their use in the community. Non-woven materials like felted cotton (cotton pad), rayon (car shop towel) or polypropylene (dust cover) may be an alternative showing high breathability and increased VLR performance with leaks present but slightly lower protection when properly sealed. Of the household materials tested the “Allison” mask, made out of two easily breathable rayon layers, may be the best design compromise since it yields intermediate VLR values in the leak scenario and *>* 90 % in all cases except dry breath when properly sealed.

The weakly electret materials (in blue) have VLR values in between those of household materials and N95 respirators in accordance with their FPF (e.g. Fig. 1b). The surgical-style mask is in the middle of the spectrum reducing viral load by 50 % with leaks and nearly 100 % when sealed. Halyard H600 shows the same filtration performance when sealed but decreased VLR values when leaks are present due to its high pressure drop. Filtrete and Blue Air filters are at the upper end of the VLR spectrum but it may be challenging to make masks from these thin materials. Their test results, however, are encouraging indicating effective air filtration in enclosed spaces is possible with such filters.

Our KN95 masks (in yellow) perform poorly with leaks present compared to N95 respirators because their *A*_M_ is much smaller (1/6 of a N95 respirator), which increases flow resistance pushing more unfiltered air through the leak. The small *A*_M_ is likely an underestimate for some KN95 models but is intended as worst-case scenario since the ones tested here deflate during inhalation. The dramatic improvement of KN95 performance with increasing *A*_M_ is highlighted in Fig. 3a for dry breath with leaks present showing VLR values in excess of those observed for N95 respirators at comparable *A*_M_. A similar effect is observed for the Allison mask (Fig. 3b) so a good mask design should aim to maximize surface area. The increase of VLR with increasing *A*_M_ is small under leak-free conditions (bottom row) because it uniquely depends on FPF changes with flow rate in this case. VLR is also a function of breathing rate but VLR increases with increasing minute volume are small compared to the changes induced by varying *A*_M_.

**Fig. 3.**
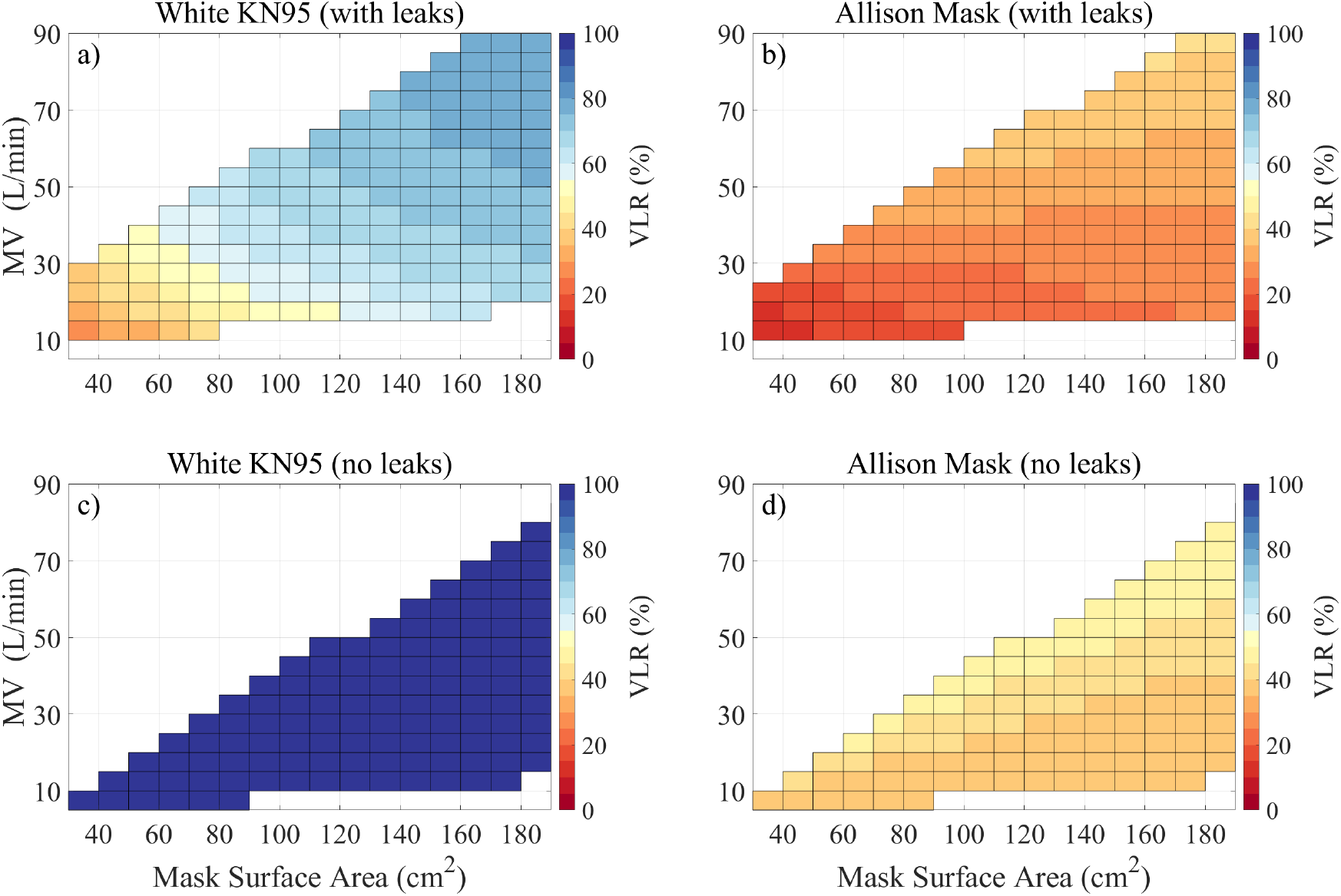
Viral load reduction as function of face mask surface area and minute volume. Leak conditions for breath aerosol at dry conditions (50 % RH) are shown in the top row, while bottom plots show model results without leaks. Left and right columns depict VLR dependency (coloration) for white KN95 and Allison mask, respectively. Uncolored regions are outside the measured flow speeds.

In summary, our model shows that the unexpected efficiency of face masks in reducing viral aerosol transmissions is the synergy of two effects: their excellent filtration performance above 1 *µ*m and the fact that the majority of exhaled aerosol volume is found above this diameter. In other words, face masks let a large portion of exhaled aerosols through because most of these are smaller than 1 *µ*m in diameter, but block most of the viral load, which is carried on larger particles. A particle with a diameter of 3 *µ*m has 1000 times the viral load of a 0.3 *µ*m aerosol so is far more important to filter. While this means the nearly complete protection required in hospitals is only achieved with a properly fitted N95 respirator, it also reinforces the importance of wearing face masks in public during a pandemic. Our calculations are intentionally pessimistic; the small surface area, big leaks, and gentle breathing rate are all reasonable, but bias VLR downward, so these masks may perform better when worn properly.

## Data Availability

Data and code will be made available online upon publication in a peer-reviewed journal.

## Acknowledgments

This work was supported by NSF grant NNX15AE44G, which was not intended for this but used in response to the worldwide COVID19 pandemic. We appreciate the help of Ian Wasnich who did initial breathing efficiency tests of household materials at Oceanit laboratory. We are grateful to the Hawaiian designers and face mask makers Jana Lam, Allison Izu, Angel Yanagihara and Sergey Negrashov and would especially like to thank Gabi Weiss for bringing researchers and mask makers together. We thank Antony Clarke for helpful comments.

## Methods

### Experiment and instrumentation

The Hawaii Group for Environmental Aerosol Research (HiGEAR) instrumentation suite is repurposed for this work to challenge a variety of proposed alternative mask materials with sodium chloride (NaCl) aerosol in a manner similar to the NIOSH certification procedures for respirators. Test aerosol is generated by nebulizing a solution of NaCl (0.02 g ml^*−*1^) and ultra pure deionized water with a particle generator (B&F Medical Supplies, LLC) then adding desiccated filtered air to reduce RH past the 45 % efflorescence point of NaCl. All flows are regulated by flow controllers (Alicat Scientific) to ensure a stable aerosol concentration during experiments. This produces a monomodal log-normal particle size distribution with a geometric mean diameter of 0.115 *µ*m and a standard deviation of 2.27. The dried particles are then diluted in a mixing chamber with filtered room air to a concentration of 1100 cm^*−*3^ (see flow chart in Extended Data Fig 1).

The aerosol passes through a ^210^Po-based charge neutralizer (a custom housing with 3 Staticmaster 2U500, NRD), before it reaches the sample material clamped into a filter holder made from NW 25 ISO-KF hose adapters (MKS Instruments; exposed area of the filter sample is 6.88 cm^2^). This neutralization of test aerosol is an extremely important step because air friction charges test aerosols during the nebulization process ^46^. If left untreated, charged test aerosol can artificially enhance filtration efficiency of test materials because it induces charge separation in oncoming particles (dielectric effect) once trapped on the material fibers. As pointed out by Ou et al. ^42^ this may have to be considered as limiting factor when interpreting recent studies that show unusually high filtration efficiencies of household materials in the submicrometer aerosol range ^47^.

NaCl aerosol number concentration is obtained with condensation nuclei counters (0.01 - 3 *µ*m, CN 3010, TSI) up- and downstream of the filter holder during the experiment. Particle size distributions are measured with 3 instruments: a long Differential Mobility Analyzer (0.01 - 0.5 *µ*m, LDMA 3071A, TSI), an Ultra High Sensitivity Aerosol Spectrometer (0.06 - 1 *µ*m, UHSAS, DMT) and an Aerodynamic Particle Sizer (0.5 - 10 *µ*m, APS 3321, TSI). The UHSAS has been shown to agree well with the LDMA for NaCl aerosol ^48^. APS aerodynamic diameters are converted to geometric diameter using the dry NaCl density of 2.16 g cm^*−*3^. For low flow rate tests, the APS was replumbed to recycle the 4 L min^*−*1^ sheath flow from the exhaust, reducing its inlet flow to 1 L min^*−*1^.

### Testing flow speeds

Material tests are conducted at 3 flow speeds (2.7 cm s^*−*1^, 8 cm s^*−*1^ and 22.7 cm s^*−*1^). The NIOSH N95 respirator testing protocol uses a flow speed of 14.7 cm s^*−*1^, but it is useful to have a range of flow rates to account for variable surface areas of homemade masks as well as breathing rates or minute volumes (*MV* ; inhaled air per minute). Assuming breathing can be divided equally into 3 parts (inhalation, holding breath, exhalation) mask surface area *A*_M_ and *MV* are related to mask flow speed *v*_M_ via

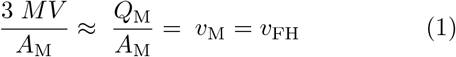

where *Q*_M_ is the mask flow rate and *v*_FH_ the velocity through the filter holder. Typical hemispheric N95 8210 respirators have a *A*_M_ of 189 cm^2^ as measured earlier ^44^, so our 3 flow speeds yield *MV* estimates of 10, 30 and 86 L min^*−*1^, which correspond to resting, moderate and strenuous activity levels ^45^, respectively.

### Filter penetration fraction

The UHSAS switches between up- and downstream aerosol with samples in place, while the higher flow rate LDMA and APS measurements are obtained with and without mask material in the filter holder. Filter penetration fraction for each flow setting is then defined as the ratio of average downstream (*D*_d_) to upstream (*D*_u_) size distributions, corrected for any changes in nebulizer output by the ratio of upstream CN concentrations during the upstream (*UCN* _u_) and downstream (*UCN* _d_) periods:

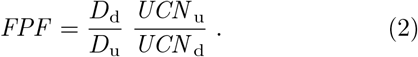

FPF over the complete size range tested (0.02 - 3 *µ*m) is obtained by joining measurements from individual instruments. Overlapping instrument data are averaged.

Since FPF is a ratio, instrument errors, counting efficiency issues and flow-dependent tubing losses cancel each other when calculating measurement uncertainty so the error range ΔFPF depends mainly on the stability of the aerosol generation over time. Since equation 2 accounts for overall concentration changes, it is the shape of the distribution that must be evaluated. Nebulizer variability is certainly the same during upstream and downstream periods, so we use the size-dependent fractional standard deviation *ε* (standard deviation/mean) of the upstream distributions (*D*_u_*/UCN*_u_) over the course of the whole experiment. The 1*σ* estimated error range is then and is shown as shading in Figure 1 and Extended Data Figures 2 and 4. Fractional errors are large at the ends of the data because the nebulizer produces few particles at those sizes.

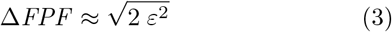

Sample to sample variability for an N95 mask is shown in Extended Data Fig. 3 for three cuts from a single 3M 8210 model. Differences in sample penetration fraction are about 5 % at the high velocity setting (in red), which amounts to an additional measurement error of about 20 %, consistent with earlier research ^49^. Other materials likely feature similar inhomogeneities but we do not have sufficient samples to quantify that.

### Air flow resistance

Air flow resistance *R*_Flow_ or breathability is calculated by measuring pressure drop Δ*p* across materials using the internal pressure sensor of the UHSAS unit (replumbed to measure pressure in the sample line). The effect on pressure readings induced by varying tubing length between up- and downstream setting is accounted for by assessing pressure drop Δ*p*_t_ at a given *v*_FH_ with no material in the filter holder. *R*_Flow_ then becomes

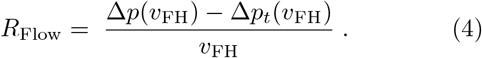

Extended Data Table 1 lists *R*_Flow_ as a velocity-independent material constant by averaging medium and high flow settings. This can be done because Δ*p* linearly increases with *v*_FH_. Low flow tests are ignored since pressure differences are small compared to the measurement uncertainty of the UHSAS pressure sensor.

### Multiple material layers

The effect of multiple layers on FPF was evaluated for Halyard H600 (Extended Data Fig. 4) and Filtrete 1500 (not shown) because the simple FPF^*n*^ formula that applies for *n* layers of a non-electret filter material ^38^ (e.g. 100 particles filtered at FPF of 0.6 yield 60 after the first layer and if filtered again 36 particles) has to be weighted by uncharged and charged aerosol fraction. Air flow resistance for multiple layers of the same material is additive for both non-electret and electret materials.

### Existing observations of exhaled aerosol

Reconciling existing publications about the size distributions of exhaled aerosol is not straightforward. There are no widely accepted protocols (deep or shallow breathing, type of vocalization, methods of collecting aerosol). Different types of instrumentation are used, and each has limitations on accuracy, size range, and even definition of diameter. Moreover, person-to-person variability appears to be quite large, up to 3 orders of magnitude in aerosol concentration. Nevertheless, as discussed here we find enough common ground between studies that reasonable size distributions can be derived and mask performance can be evaluated.

Available breath and speech aerosol observations are shown in Extended Data Fig. 5. Aerosol data above 10 *µ*m in diameter are ignored as these are not the focus of this study. Data are either directly taken from manuscripts ^14,16,17^ or extracted from figures ^13,15,18,19^. Measurements made with a Grimm (Model 1.108, Grimm Aerosol Technik, Ainring, Germany) optical particle counter (OPC) are corrected by us ^14^ or by the authors ^15,18^ according to Holmgren et al. ^15^, who account for refractive index differences between exhaled aerosol and polystyrene latex calibration spheres. Other OPC data ^13,16^ are used as is since no correction is easily available. Measurements made with an APS ^17,19^ are corrected from aerodynamic to geometric diameter by dividing by the square root of the density of an aqueous NaCl solution at reported RH. The correction decreases APS diameters by about 11 %. Observations made with a Differential mobility analyzer (DMA) ^15^ are not corrected further. All but one study provide respiratory aerosol measurements as number concentration over size. Data from Larsson et al. ^18^ are shown as aerosol mass and are converted to number concentration using the density provided (1 g ml^*−*1^).

It is useful to fit aerosol data to log-normal modes, defined as ^50^

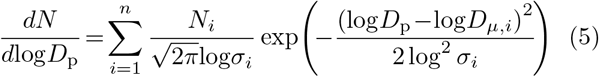

which describe total particle number concentration *N* per log_10_ unit of diameter *D*_p_ as the sum of *n* log-normal particle modes, each of which is defined by its mode number concentration *N*_*i*_, geometric mode mean diameter *D*_*µ,i*_ and geometric standard deviation *σ*_*i*_. The three parameters for each mode are derived with an unconstrained least squares minimization scheme that uses the number, area and volume distributions simultaneously for studies where we fit two modes ^13^,19. We use volume distributions only where we fit a single mode ^14^,18. OPC and DMA data from Holmgren et al. ^15^ are sufficiently inconsistent that a least squares fit cannot match the data very well (as pointed out by the authors) so we fit the data visually to agree with measurements.

Log-normal fit parameters for each data set are summarized in Extended Data Table 2. Schwarz et al. ^16^ and Johnson et al. ^17^ provided log-normal fits; we calculated the others. At just 0.07 µm, lung mode 1 is visible only with a DMA ^15^, while lung mode 2 can be measured with an OPC or DMA. The larynx mode can be observed by both APS and OPC but most studies did not provide data for the larger OPC size bins, presumably because number concentrations were very low.

Quiet breathing appears to produce as many as 3 modes of particles, although most studies only show one clearly (Extended Data Fig. 5 and Extended Data Table 2). All of the studies show a mode centered at 0.28 - 0.39 µm (though the number concentrations vary by over 3 orders of magnitude). Much of lung mode 1 is smaller than the virus and it has no more than 15 % of the aerosol volume, so is ignored here. There are so few particles in the larynx mode that only one study had sufficient data to define it at 1.2 µm, though there are hints of such a mode in others (Extended Data Fig. 5). With such limited data we cannot include this larynx mode in the “breath aerosol” size distribution, even though it may well include twice the volume of the smaller modes. That means our estimates of viral load reduction are pessimistic (biased low), since these relatively large particles are effectively filtered by all of the materials tested.

Aerosol emitted while vocalizing (“speech aerosol”) appears to have similar size modes as quiet breathing, but the larynx mode is much more prominent. We include both modes of this speech aerosol, although the volume of lung 2 mode is inconsequential at only 0.65 % of the larynx mode (Extended Data Fig. 6). Our speech aerosol distributions only include data from work using APSs, since the one study with an OPC provides insufficient data to resolve the two modes. It is not clear whether the larger lung mode 2 diameters found by the APS are real or a product of experimental differences (uncertain RH and quickly dropping APS detection efficiency below 0.9 *µ*m ^17,24^). However, these differences matter little when only considering aerosol volume as illustrated in the bottom row of Extended Data Fig. 5 for APS measurements from Asadi et al. ^19^. By replacing the log-normal fits of lung mode 2 with forced fits (*D*_*µ*_ = 0.37 *µ*m and *σ* = 1.6) that resemble fit parameters for breathing, we show both fit types yield almost identical aerosol volume distributions.

The actual concentration of aerosols and viruses is in general unknown, but we are interested in the fraction removed, so only the relative concentrations as a function of diameter are relevant. Therefore the span of almost 3 orders of magnitude in reported number concentration or the lack of a reported concentration ^16^ does not affect our analysis.

An important source of difference between studies is the relative humidity at the point of measurement. After exhalation, the concentration of water in particles is a strong function of RH, particularly at the high humidities at which aerosols are exhaled. Diameter, density, and refractive index are all dependent on the water concentration, so even small deviations from exhalation RH change particles significantly (and it is difficult to accurately control very high humidities). One can make more precise diameter measurements after drying the aerosols, but then the initial diameter must be calculated. Unlike some studies that used an RH-independent correction factor *ψ* to obtain the initial diameter ^17,51^, here we use electrodynamic balance studies on NaCl particles ^52^ to determine changes in diameter as a function of RH (similar to an earlier report ^53^) because respiratory fluids can be modeled as an aqueous NaCl solution with a water activity (equivalent to RH) of 0.995^53^.

We convert our log-fit parameters from reported measurement humidity (when reported, otherwise we calculate RH from measurement temperature, assuming breath was exhaled at 34 °C ^15^,54) to 99.5 % and 50 % RH by dividing geometric mode mean diameters by *ψ*(*RH*) (Extended Data Table 2). The humid condition is for particles exhaled through the mask, representing the extent to which the mask wearer is preventing spread of the virus. “Dry” conditions show protection offered the wearer to inhalation of breath and speech aerosol expelled by another person. The humidity-adjusted fit parameters are converted to aerosol volume then averaged. These volume fit parameters for breath and speech aerosol are summarized in Extended Data Table 3 and are shown together with the distributions used in the averaging in Extended Data Fig. 6. We set breath aerosol volume to an arbitrary value of 1 µm^3^cm^*−*3^ since we only consider lung mode 2.

### The road to viral load

A key assumption we make is that virus concentration in exhaled aerosol is equal to that in the respiratory fluids from which the particles form. That means the amount of virus on each particle is proportional to its volume and the overall exhaled viral load *VL*_T_ is proportional to the integral of the aerosol volume distribution, which is easily obtained by converting the number concentration size distribution assuming particles are spherical (see equation 5 and Extended Data Table 2). The viral load penetrating the face mask *VL*_M_ is proportional to the integral of the product of the volume size distribution and filter penetration fraction (see bottom row of Fig. 1). In the absence of leaks, the viral load reduction (expressed as a percentage) is then simply the ratio of these two viral loads:

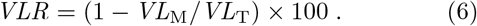

Filter penetration fraction varies considerably with air velocity (Extended Data Fig. 4) so is included in the calculation of *VL*_M_ by interpolating between FPF data made at the 3 flow speeds. FPF values above 3 *µ*m are zero except for the coffee filter and dust cover fabric, which have been extrapolated beyond 3 *µ*m. The test velocities *v*_M_ that are used to create Fig. 2 are calculated via equation 1 based on a *MV* of 10 L min^*−*1^ (breathing at rest) and a surface area of 189 cm^2^ for N95 respirators ^44^ and 31 cm^2^ for all other face masks (assuming a spherical cap shape with a diameter and height of 3 cm and 1 cm).

### Modeling face mask leakage

To estimate VLR under more realistic conditions, with poorly fitted masks that leak on either side of the nose, we model leak paths as a pair of tubes 3 cm long and 1 cm in diameter. Some of the virus-laden air still goes through the mask, but some fraction will divert through the leaks and remain unfiltered. VLR including the leak then becomes

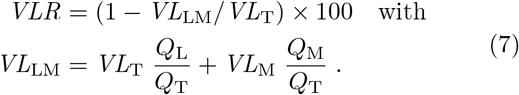

where *Q*_L_, *Q*_M_, and *Q*_T_ = *Q*_L_ + *Q*_M_ are flows through the leak, the mask and the total exhalation (or inhalation) rates, respectively. Note, this relationship simplifies to equation 6 if leaks are eliminated (*Q*_L_ = 0). The flow rates *Q*_L_ and *Q*_M_ are not immediately known but must satisfy the requirement that the pressure drops across face mask Δ*p*_M_ (see Eqs. 1 and 4) and leak Δ*p*_L_ are equal. This condition is described by a set of nonlinear equations, which we iteratively solve for *Q*_L_ and Δ*p*:

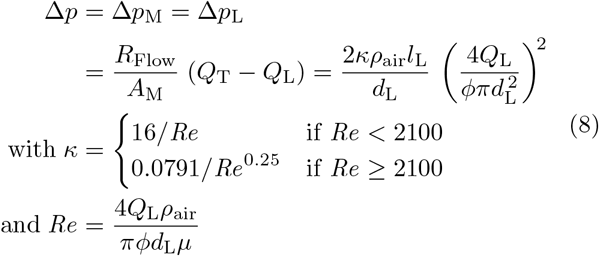

with leak length *l*_L_ and diameter *d*_L_, air density *ρ*_air_ (at 298.15 K and 1013.25 hPa), number of leaks *φ*, air viscosity *µ*, and the flow Reynolds number *Re*, which indicates laminar flow when below 2100 (Δ*p*_L_ via equations 6.1.4, 6.2.11 and 6.2.12 in chapter 6 of Bird et al. ^55^). Our leak model reproduces lab measurements of the total inward leakage of NaCl aerosol ^39^, defined as the sum of aerosols that make it through a N95 respirator as well as leaks, to within 10 % when we input the study’s test aerosol distribution and the dimensions of the tubes that were used as leak paths through a N95 respirator. We assume their N95 respirator has the same filtration efficiency signatures and pressure drop as the N95 8210 tested here. While only an estimate, this provides an indication that our leak calculations are reasonable.

Our choice of a small surface area for all face masks (*A*_M_= 31 cm^2^) but the two N95 respirators assumes that only a small region around the nose and mouth are available while inhaling, as many masks tend to collapse against the face. This is likely to be an underestimate during exhaling, but the leaks are expected to be smaller too, and both vary with face contours, mask design, and fit in ways that are difficult to model. The big leak, small filter area, low minute volume, and limitation to lung mode 2 in Fig. 2 all bias the VLRs down; this is intended as a worst-case scenario with ill-fitted masks. If efforts are made to reduce leaks and increase filtration area, protection is substantially increased, even with non-electret masks, as shown in Fig. 3 for two selected face masks.

A detailed error analysis for VLR is not practical because instrument and measurement errors of existing breath and speech aerosol studies are not clear, and we do not know whether the viruses partition uniformly into aerosol volume. VLR uncertainty is, however, expected to be small in comparison to changes induced by RH, *Q*_T_, *A*_M_ and of course leak size, as discussed in the manuscript.

---

**Extended Data Fig. 1.**
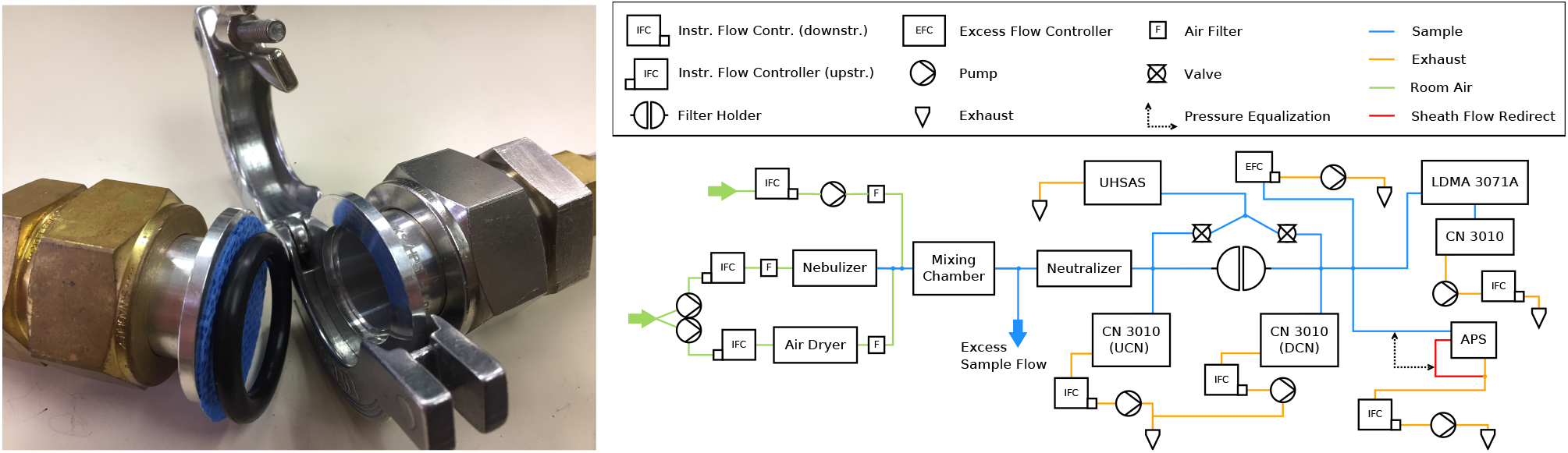
Instrument setup. Aluminum hose adapters used as filter holder with a single layer of Halyard H600 (left) and flow schematic for sodium chloride aerosol tests (right).

**Extended Data Table 1.** Description and air flow resistance of tested filter materials. Respirators and masks are separated from electret and fabric materials not intended for face mask usage by manufacturer. All polypropylenes (PP) are non-woven.

| Name | $R_{\text{Flow}}$ (hPa s m<sup>-1</sup>) | Material Description |
| --- | --- | --- |
| Berkshire Mask | 5.78 | Berkshire BCR® BM.1.12 3-layer mask (PP) |
| N95 8210 | 9.06 | 3M® N95 8210 respirator (PP) |
| N95 8000 | 11.59 | 3M® N95 8000 respirator (PP) |
| KN95 (black) | 10.25 | Black outer layer with sewn in mask (Tengfei Tech.) |
| KN95 (white) | 20.37 | White mask (online purchase; manufacturer unknown) |
| Filtrete 2500 | 1.45 | 3M® Filtrete™ 2500 air filter (PP) |
| Filtrete 1500 (1L) | 1.46 | 3M® Filtrete™ 1500 air filter (PP) |
| Blue Air Filter | 1.73 | Blue Air® HEPA single layer filter (PP) |
| Filtrete 1500 (2L) | 2.93 | 3M® Filtrete™ 1500 air filter (PP) |
| Filtrete 1500 (4L) | 5.86 | 3M® Filtrete™ 1500 air filter (PP) |
| Halyard H600 (1L-w ) | 9.15 | Autoclaved Halyard® H600 (white layer only) |
| Halyard H600 (1L-b) | 12.70 | Autoclaved Halyard® H600 (blue layer only) |
| Halyard H600 (2L-bw) | 21.78 | Autoclaved Halyard® H600 (2-ply; spun; PP) |
| Dust Cover Fabric | 2.75 | 1 layer of washed CanvasETC® dust cover (PP) |
| Car Shop Towel | 3.97 | Flinka™ shammy towel for car (100% Rayon) |
| Cotton Pad | 4.92 | Felted pad (100% cotton; 3 mm thick) |
| Allison Mask | 14.25 | 1L 92% Rayon 8% Spandex & 1L Ponte de Roma (400 g m<sup>-2</sup>) |
| Paper Towel & Kleenex | 14.65 | 1 Kleenex® between 2 True Home CVS® towels |
| Coffee Filter (2L) | 18.18 | 2 CVS® coffee filter layers (disposable; basket-style) |
| Angel Mask | 23.05 | 2 layers Pima (150 TPI) and 2 layers Quilter's cotton (64 TPI) |
| Serge Mask | 23.68 | 2 layers of loosely woven cotton sheet (71 TPI) |
| Jana Mask | 55.84 | 1 layer of tightly woven cotton fabric (58 TPI) |

**Extended Data Fig. 2.**
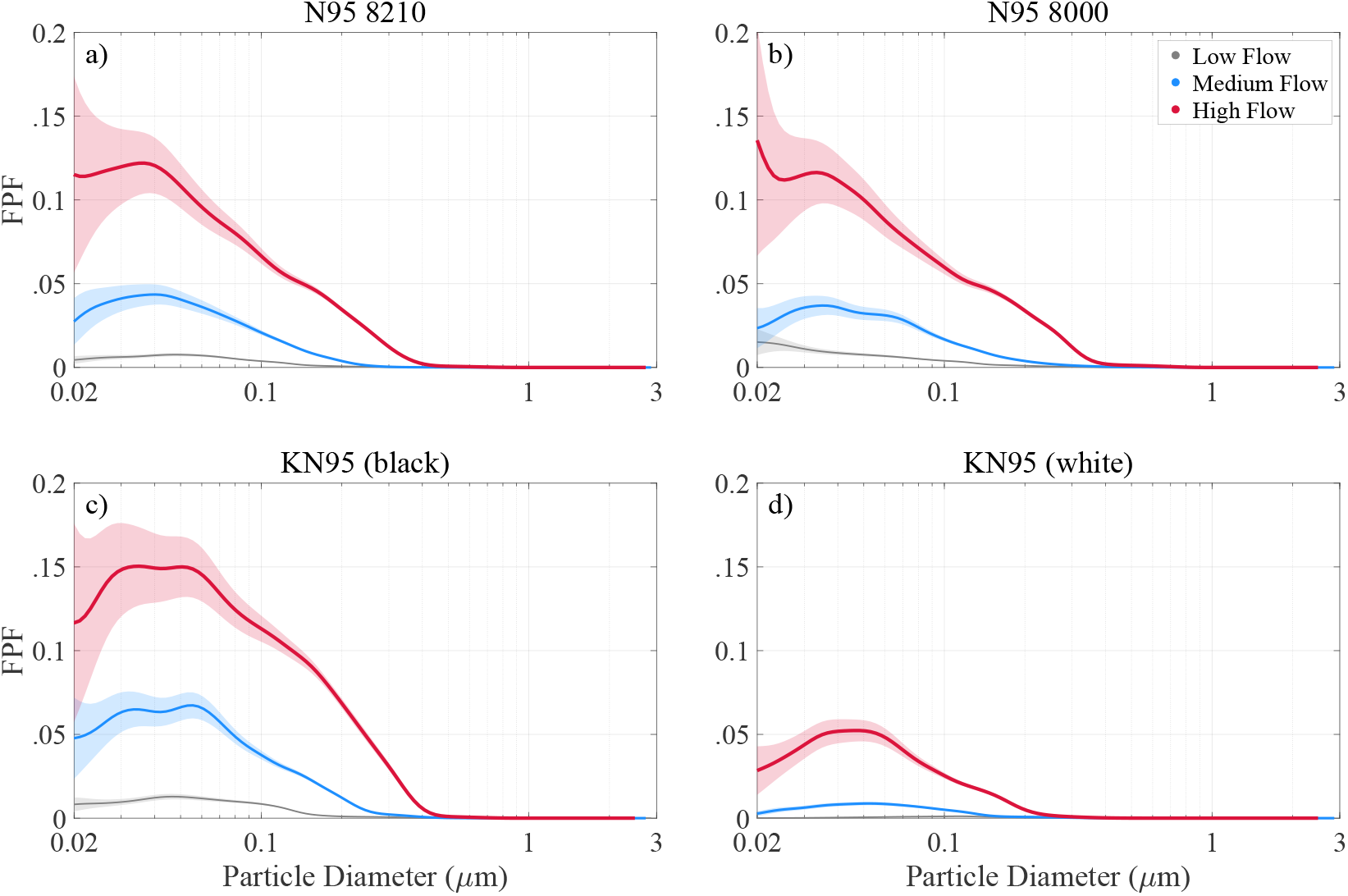
Filter penetration fraction for selected N95 standards. Observations at low (2.7 cm s^*−*1^), medium (8 cm s^*−*1^) and high flow (22.8 cm s^*−*1^) are shown in gray, blue and red, respectively. Shading represents measurement uncertainty (1*σ*) around the mean (solid lines).

**Extended Data Fig. 3.**
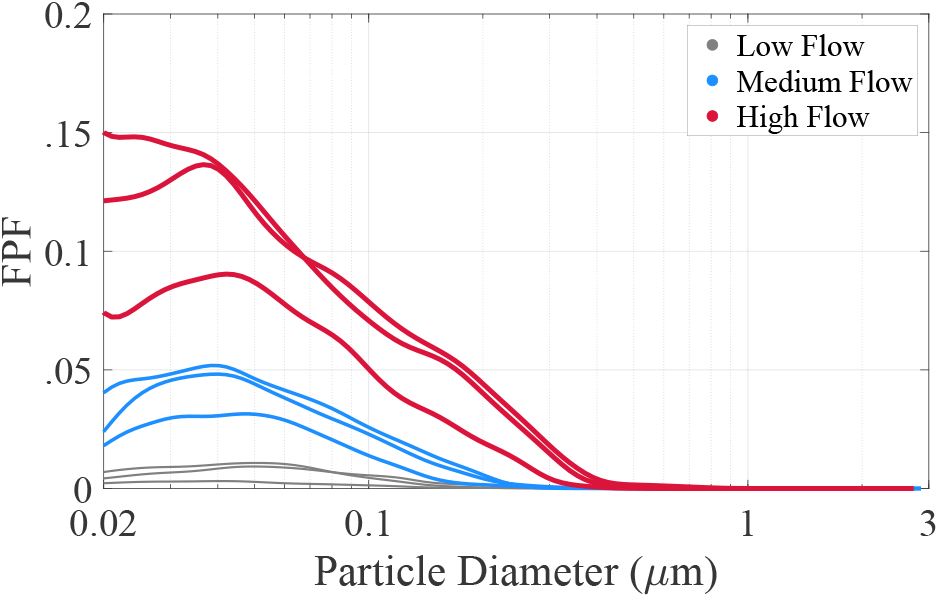
Filter penetration fraction of 3 different samples cut from a single N95 8210 respirator. Solid lines represent the mean at low (2.7 cm s^*−*1^), medium (8 cm s^*−*1^) and high flow (22.8 cm s^*−*1^) in gray, blue and red, respectively. Measurement uncertainty is omitted for clarity.

**Extended Data Fig. 4.**
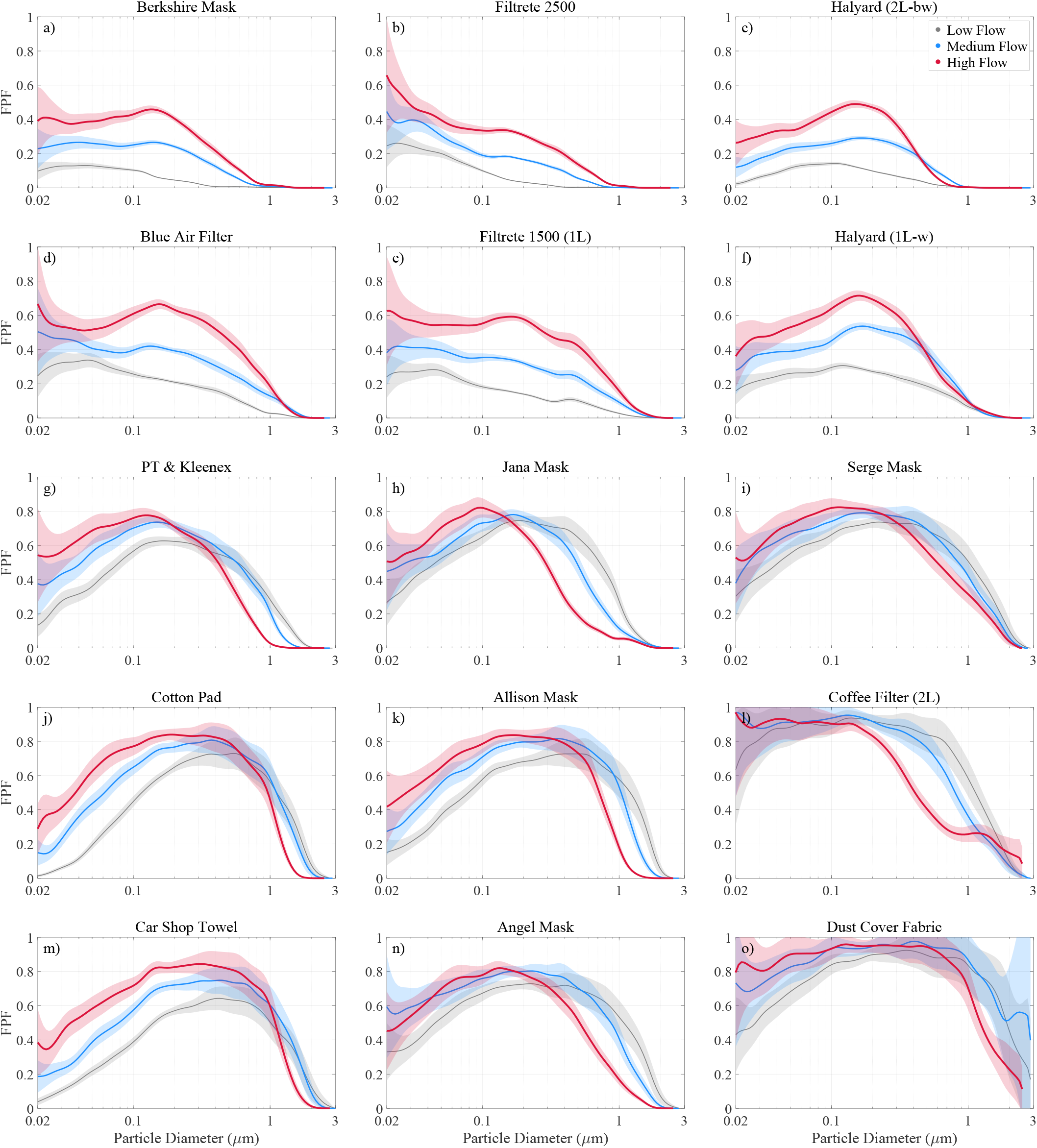
Filter penetration fraction for various materials. Observations at low (2.7 cm s^*−*1^), medium (8 cm s^*−*1^) and high flow (22.8 cm s^*−*1^) are shown1 in gray, blue and red, respectively. Shading represents measurement uncertainty (1*σ*) around the mean (solid lines).

**Extended Data Fig. 5.**
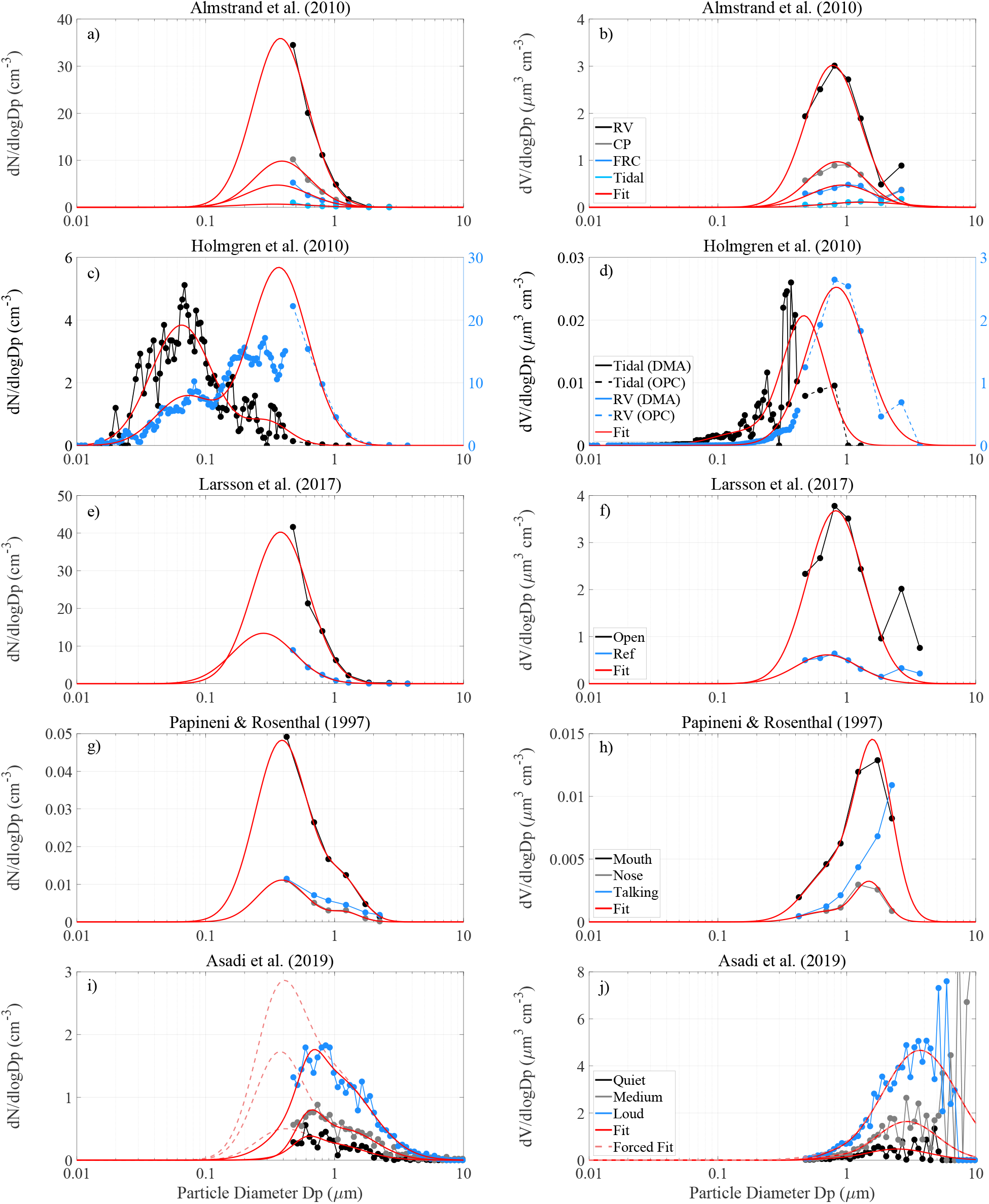
Log-normal fits through existing breath and speech aerosol data. Left and right columns show log-normal number concentration (*dN/d*log*D*_p_) and volume size distributions (*dV/d*log*D*_p_) superimposed onto observations sorted by publication, respectivel1y. Log-normal fits derived with least squares minimization scheme are shown as solid red lines, while fits with fixed mode parameters (*D*_*µ*_ = 0.37 *µ*m and *σ* = 1.6) are shown as dashed red lines (bottom row only).

**Extended Data Table 2.**
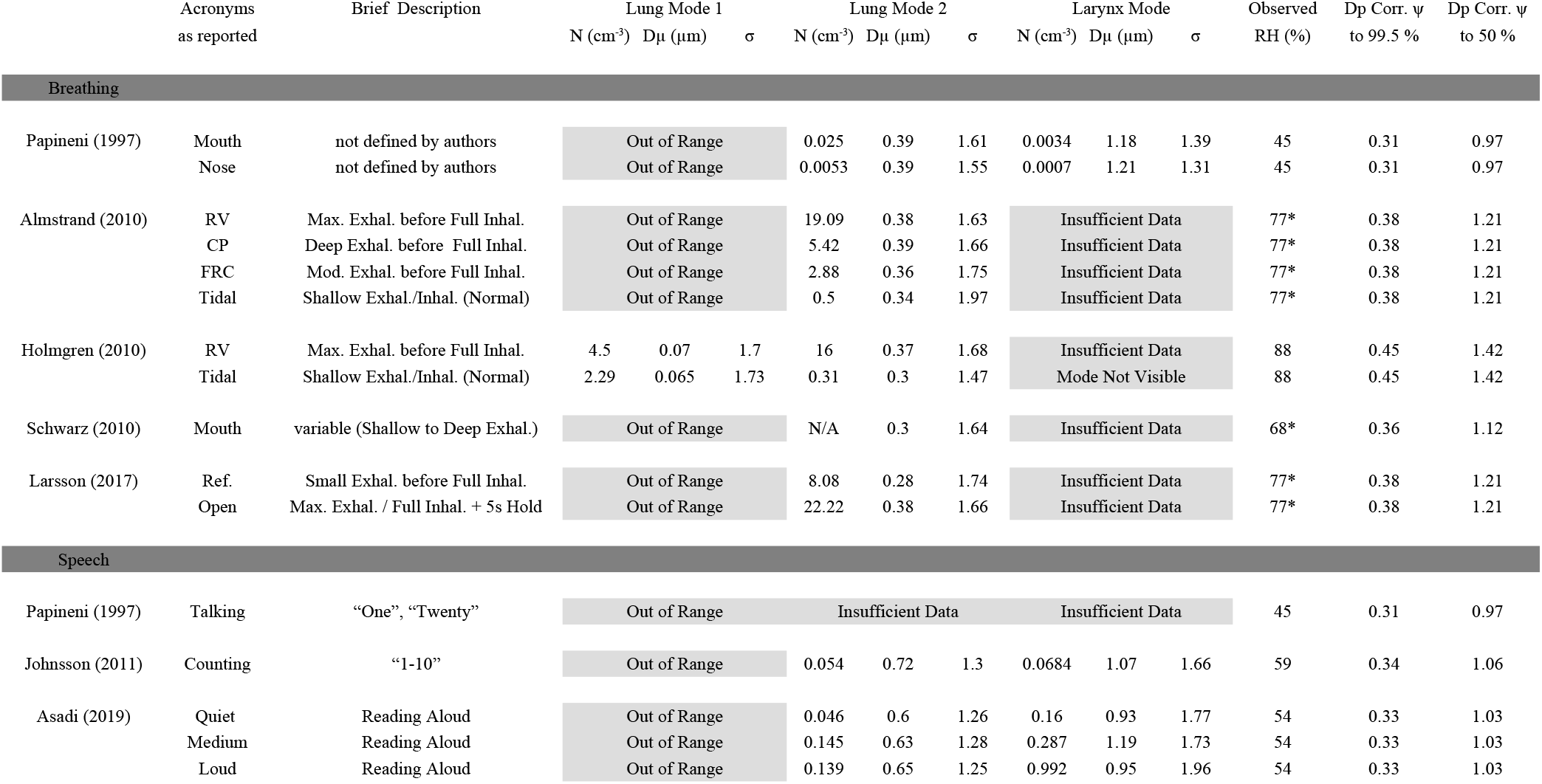
Summary of existing breath and speech aerosol observations. Acronyms listed are as used by authors. Measurements have been adapted from studies and are represented by a log-normal fit consisting of up to two modes. Each aerosol mode can be expressed by total number concentration *N*, the geometric mode mean diameter *D*_*µ*_, and its geometric standard deviation *σ*. Corrections provided in the last two columns adjust *D*_*µ*_ at observation relative humidity (RH) to humid (99.5 %) and dry (50 %) conditions. The humid condition applies during exhalation through the face mask, while dry conditions represent aerosol inhalation (expelled by another person) at a moderate ambient RH. Observation humidity shown is either directly provided by the authors or calculated via temperature of measurement and exhaled air (assuming exhaled air is at 34 °C; marked with *\**).

**Extended Data Table 3.** Aerosol volume fit parameters. Mean breath and speech aerosol volume log-normal fit parameters derived from existing observations at selected relative humidity (RH). Breath volume only considers lung mode 2 with mode volume arbitrarily set to 1 *µ*m^3^cm^*−*3^ (see methods), while speech volume is derived from lung mode 2 and larynx mode.

| Volume Type | RH (%) | $V$ (μm<sup>3</sup>cm<sup>-3</sup>) | $D_\mu$ (μm) | $\sigma$ | $V$ (μm<sup>3</sup>cm<sup>-3</sup>) | $D_\mu$ (μm) | $\sigma$ |
| --- | --- | --- | --- | --- | --- | --- | --- |
| Breath | 50 | set to 1 | 0.68 | 1.67 | - | - | - |
|  | 99.5 | set to 1 | 2.15 | 1.67 | - | - | - |
| Speech | 50 | 0.016 | 0.75 | 1.27 | 1.10 | 2.75 | 1.78 |
|  | 99.5 | 0.485 | 2.33 | 1.27 | 33.5 | 8.59 | 1.78 |

**Extended Data Fig. 6.**
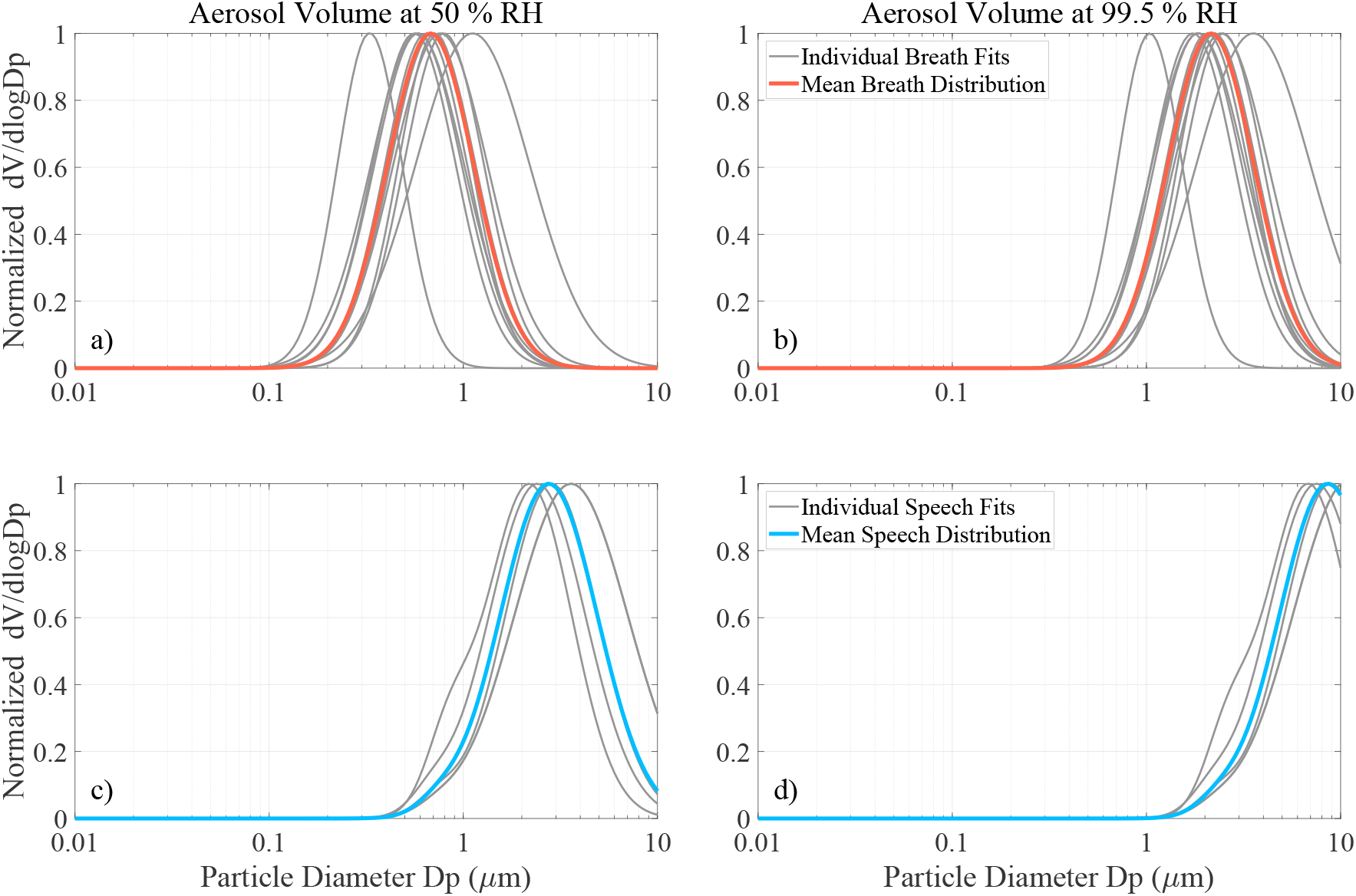
Normalized average breath and speech aerosol volume distributions. Mean breath and speech aerosol volume distributions derived from individual volume fits (in gray) are highlighted in black and blue, respectively. Mean volume fit parameters are summarized in Extended Table 3 and are calculated from humidity-adjusted number concentration log-normal fit parameters (provided in Extended Table 2). The data are normalized to the maximum value to illustrate distribution shape. Aerosol volume in the left column approximates inhalation of aerosol expelled by another person, while humid conditions represent exhalation through the face mask.

